# The day-to-day experiences of caring for children with Osteogenesis Imperfecta: A qualitative descriptive study

**DOI:** 10.1101/19007187

**Authors:** Aimee R. Castro, Jessica Marinello, Khadidja Chougui, Marilyn Morand, Claudette Bilodeau, Frank Rauch, Argerie Tsimicalis

## Abstract

**Aims and objectives:** This study aimed to explore the day-to-day experiences of caregivers who are caring for children with Osteogenesis Imperfecta (OI).

**Background:** OI is a rare genetic condition known to cause bone fragility. Family caregivers, such as parents, of children with OI play an important role in helping these children live well at home.

**Design:** The design was qualitative description.

**Methods:** A qualitative descriptive study was conducted which adheres to the COREQ guidelines. Adult caregivers (n=18) of children with OI were recruited at a children’s hospital in Montréal, Canada to participate in individual interviews. The interviews were transcribed verbatim and inductively thematically analysed.

**Results:** The following caregiving themes were identified in these interviews: regular day-to-day caregiving activities, including morning routines, evening routines, and the facilitation of their child’s mobilization; periods that made the caregiving routine more challenging, such as fractures, surgeries, and pain; and the long-term strategies caregivers developed to support day-to-day care, such as managing the environment, accessing medical and school resources, and coordinating care and respite.

**Conclusions:** The results showcase what being a caregiver for a child with OI involves on a day-to-day basis.

**Relevance to clinical practice:** The recommendations include suggestions for future clinical, policy, and research endeavours to develop better policies and interventions to support the unique needs of family caregivers of children with OI. These recommendations may be relevant to other clinicians and policymakers working with families living with rare and chronic physical conditions.

## Introduction

Osteogenesis Imperfecta (OI) is a rare genetic condition affecting 1 in 10 000 births that is predominantly caused by an alteration in the genes responsible for collagen production (Marini et al., 2017; Trejo & Rauch, 2016). It is characterized by increased bone fragility, but this chronic condition can also cause teeth and soft tissue abnormalities, discoloured sclera, hearing loss, shortened stature, and skeletal deformities (Marini et al., 2017; Trejo & Rauch, 2016). OI Types I – IV are the original classifications of OI, although other types have been added since (Trejo & Rauch, 2016). OI Type I is the most common form of the condition and has the mildest disease severity, with fewer fractures expected over the life course. OI Type II is lethal in the neonatal period. OI Type III is the most severe form of OI that is compatible with a longer lifespan. OI Type IV is of moderate disease severity. OI Types V, VI, and VII are newer classifications that present with different tissue and bone mineralization phenotypes, and have mild to moderate severity (Marini et al., 2017; Trejo & Rauch, 2016). At present, there is no cure for OI. Bisphosphonate therapy is used to improve bone mass density (Trejo & Rauch, 2016). Surgery, physical therapy, and occupational therapy are other common treatment modalities (Dogba et al., 2013; Marini et al., 2017).

Family caregivers, such as parents, play a critical role in helping children with unique needs to thrive at home in the community (McCann, Bull, & Winzenberg, 2012). It is important to understand the day-to-day experiences of caregivers of children with unique needs - such as the unique care needs of children who have fragile bones - in order to develop policies and services that are adapted to their specialized daily care needs (McCann et al., 2012; Pearlin, Mullan, Semple, & Skaff, 1990). Thus, the aim of this research was to better understand the day-to-day experiences of caring for children with OI. These results have important implications for clinicians and policymakers who work with families living with OI or other rare, chronic conditions.

## Background

When a child has OI, a parent usually adopts the role of primary caregiver to assist with their child’s everyday needs (Vanz, Felix, da Rocha, & Schwartz, 2015). Challenges unique to OI caregiving include the constant risk of fractures, over-protectionism, social isolation, home adaptations, and suspicions of child abuse (Arabaci et al., 2015; Bernehall & Brodin, 2002; De Carmoy, 2004; Dogba, Rauch, Tre, Glorieux, & Bedos, 2014). OI parents have reported lower scores in the environmental quality of life domain (i.e., the domain which includes factors such as environmental safety and access to social supports) compared to the general population of parents (Szczepaniak-Kubat, Kurnatowska, Jakubowska-Pietkiewicz, & Chlebna-Sokol, 2012; Vanz et al., 2015). The OI caregiver experience may generate positive experiences for families, such as the development of an optimistic mindset and being inspired by their children’s resilience (Dogba et al., 2013). However, for many families, a child’s OI diagnosis appears to be a life-altering event requiring various and ongoing parental adjustments (Arabaci et al., 2015; Dogba et al., 2013; Murphy, Christian, Caplin, & Young, 2007).

It is important to understand the day-to-day experiences of caregivers of children with unique needs like OI in order to develop services that are adapted to the specialized daily care needs of these families (McCann et al., 2012; Pearlin et al., 1990). While several studies on the OI family experience have included some analysis of the day-to-day lives of OI caregivers (Bernehall & Brodin, 2002; De Carmoy, 2004; Deatrick, Knafl, & Walsh, 1988; Dogba et al., 2013; Dogba et al., 2014; Hill, Baird, & Walters, 2014; Santos, Pires, Soares, & Barros, 2017), none of these studies focused primarily on the minutiae and routines of daily care.

Therefore, the purpose of this study was to explore the day-to-day experiences of caregivers who are caring for children with OI. We interviewed 18 adult caregivers of children with OI about their day-to-day OI caregiving experiences. They described their routine activities of day-to-day caregiving, their experiences during challenging periods of caregiving, and the long-term strategies they developed to support their child’s day-to-day care needs. These findings have important implications for clinical practice, policy, and research.

## Methods

### Design

The study methodology was qualitative description (Sandelowski, 2000). This study was part of a larger one which sought to explore the views of OI caregivers on using Internet-based technologies to support their caregiving needs (***manuscript available as a preprint. Will add citation after double-blind peer review***).

### Sample and Recruitment

A purposive sampling strategy was used to recruit family caregivers of children who were being treated for different types of OI at the study site. Inclusion criteria included all family caregivers of patients being treated for OI at the hospital site, who considered themselves to be the primary caregivers of the child, and who spoke either English or French. A non-authoritative member of the team approached clinicians who had appointments with OI patients to mediate an introduction to the study. If caregivers expressed an interest in learning more about the study, then a member of the research team described the study and informed the caregiver of the potential consequences of participating. If the caregiver provided their written consent to participate, then an interview was scheduled at the caregiver’s convenience.

### Data Collection

[***Note to editor and reviewers: To keep this section as anonymous as possible for double-blind review, we have replaced the authors’ initials with “**”. Once the paper is reviewed, we will replace these ** with the specific contributing authors’ initials.]

A demographic data survey was filled out by each participant. Individual semi-structured interviews were subsequently conducted by a member of the research team, in either English or French depending on the caregiver’s preference. The interviews were conducted by either author ** or **, at the study site or by telephone or videoconferencing. Key questions relevant to day-to-day care from the larger study’s interview guide are listed in Table 1. The interview guide was reviewed by key informant clinicians prior to the participant interviews. Interviews were audiotaped, field notes were recorded during and after the interviews, and a reflective journal was kept by author **.

**Table 1.** Key interview questions and prompts related to day-to-day care.

|  |
| --- |
| ***Main questions:*** |
| **1) Can you describe what a regular day looks like when caring for your child with OI?** |
| **2) And then can you describe a more challenging day recently?** |
| ***Prompts:*** |
| -What are some of the duties or responsibilities of caring for a child with OI? |
| -How has your caring for your child changed over time? |
| -What areas of your family's life do you feel are going well? |
| -What are some areas that you would like to see improve? |
| -What happens on a day with a bone break? What happens in the days that follow? |
| -Do you feel you know about community resources for families of children with chronic conditions? |
| -How has caregiving affected your lifestyle and schedule? Your job? Your relationships? Your personal care? |
| -How do you feel about your social support? |
| -Do you have access to extra help and/or care around the house? |

### Analysis

The surveys were analyzed using descriptive statistics. The transcribed data and field notes were uploaded into Microsoft Excel, where they were first inductively open-coded by three of the authors (**, **, and **), and then further analyzed primarily by ** with inter-coder checks by ** using (Braun & Clarke, 2006)’s six steps for thematic analysis. Interviews conducted in French were first open-coded into English by authors ** and ** who are fluent in both English and French. In frequent meetings with the principal investigator (**), the research team discussed the ongoing development of the codebook by ** and established the final themes. These codebooks and meeting notes were recorded in a research journal, as were the ongoing research reflections of **. The carefully anonymized research journal and the codebook may be made available for audit upon request.

### Ethical Considerations

Ethical approval was received from the university’s Institutional Review Board and the study site prior to recruitment. The university’s ethical review process conforms to the Canadian Tri-Council Policy Statement on the Ethical Conduct for Research Involving Humans. All participants provided written informed consent to participate in the study, with the understanding that they would not be identifiable in any presentations of the research. All participant data were anonymized.

### Rigour

Qualitative rigour was established using (Lincoln & Guba, 1985)’s four criteria of credibility, dependability, confirmability, and transferability. Credibility was enhanced by the use of reflexive journaling, the audio-taping of interviews, and the research team’s frequent meetings for peer debriefing (Polit & Beck, 2012). The dependability of this research was enhanced by careful documentation of our team discussions and decisions in a research journal, and making the anonymized research journal, and codebook available for audit upon request (Polit & Beck, 2012). The confirmability of this research is supported by our carefully documented decision trail in our research journal, our regular team debriefings, and our audit trail (Polit & Beck, 2012). The transferability of this research is supported by the team development of our codebook, inter-coder checks, and our use of thick descriptions and quotes (Polit & Beck, 2012). The Consolidated Criteria for Reporting Qualitative Research (COREQ) guided our data collection and analysis and later reporting of the research (Supplementary File 1) (Tong, Sainsbury, & Craig, 2007).

## Results

### Demographic Data

Nineteen caregivers were approached in-person, and all verbally agreed to participate. However, one caregiver was lost to follow-up. Eighteen adult caregivers from 14 families were individually interviewed. Thirteen caregivers were women and five were men. Three families resided outside of Canada. Four of the 14 families shared that they had family incomes of less than $50 000 per year, and all 18 caregivers had received at least some post-secondary education. Interviews ranged from 15 minutes to 1.5 hours in length. They resulted in 13.05 hours of audio-recordings and over 8000 spreadsheet lines of coded transcript. Their children’s OI types included I, III, IV, and VI; the more severe forms of types III and IV were the most common. Some children with OI required no mobilizing assistance, while others used a wheelchair full-time. Further demographic data are listed in Table 2.

**Table 2.** Demographic data.

| **Demographic Trait** | **<u>n (individuals, unless otherwise indicated)</u>** |
| --- | --- |
| **Individuals interviewed** | 18 |
| **Number of families represented** | 14 |
| **Language of interview (as caregiver preferred)** |  |
| English | 12 |
| French | 6 |
| **Median caregiver age in years (range)** | 37.5 (24-57) |
| **Caregiver's sex (/18 caregivers)** |  |
| Female | 13 |
| Male | 5 |
| **Marital Status (/18 caregivers)** |  |
| Married or common-law | 14 |
| Single (never married) | 2 |
| Separated or divorced | 2 |
| **Residential region (/14 families)** |  |
| Local (< 2 hours' drive from Montreal) | 7 |
| Within Quebec, not local | 1 |
| Other Canadian region | 3 |
| International | 3 |
| **Highest level of education (/18 caregivers)** |  |
| Some post-secondary (university or college) | 5 |
| Received university or college degree/diploma | 11 |
| Postgraduate | 2 |
| **Estimated family income (/14 families)** |  |
| Less than \$25 000 | 3 |
| \$25 000 - \$50 000 | 1 |
| \$50 000 - \$80 000 | 1 |
| More than \$80 000 | 7 |
| Do not know | 1 |
| Prefers not to answer | 1 |
| **Relationship to child with OI (/18 caregivers)** |  |
| Mother | 11 |
| Father | 5 |
| Other: legal guardian | 2 |
| **Child's OI type (/14 families)** |  |
| I | 3 |
| III | 4 |
| IV | 6 |
| VI | 1 |
| **Number of families with more than 1 child with OI in the household** | 3 |
| **Ages of children with OI (17 children with OI)** |  |
| Baby (0-12 months) | 2 |
| Toddler (13 months - 3 years old) | 1 |
| Pre-school (4-5 years old) | 1 |
| School-aged (6-12 years old) | 11 |
| Teenager (13-18 years old) | 2 |
| **Range in number of fractures / surgeries per lifetime in the children with OI**<br>Note: Several families did not mention the number of fractures or surgeries their children had experienced. This question was not explicitly included in our interview guide or questionnaire. | 1 - 200 fractures / 0 - 40 surgeries |
| **How would you describe the health of your child with OI right now? (/18 caregivers)** |  |
| Excellent | 5 |
| Very good | 4 |
| Good | 8 |
| Fair | 1 |
| Poor | 0 |
| **Family history of OI (/14 families)** |  |
| Yes | 3 |
| No | 11 |

Note: This table has been adapted for this report from the demographic data table portrayed in (\*\*\*\*Citation will be included after double-blind peer review\*\*\*). The two tables are similar but not identical.

### Caregiving Themes

We inductively derived the following themes relating to the day-to-day experiences of OI caregiving: regular day-to-day caregiving activities, challenging periods, and the long-term strategies caregivers developed to support day-to-day care.

#### Regular day-to-day caregiving activities

OI caregivers were asked to share what a regular OI caregiving day looked like. Key components of these day-to-day activities included following morning and evening routines to assist with their child’s activities of daily living (ADLs), and facilitating mobilization.

#### Morning and evening routines

Mornings were particularly busy for caregivers, when many ADLs tended to occur all at once, and all tasks needed to be completed before school and paid work began. Mornings consisted of waking everyone up, making lunches, helping children get dressed, and using the bathroom. Some children needed assistance in dressing with their leg braces or corsets. Children needing the most assistance with ADLs were younger children and those with more severe types of OI. Chronic pain management was also a part of the morning routine for at least two families. Most families chose to drive their children to school, although one child used an adapted bus. One mother said her son would walk to school if it was nice outside, but if his backpack was heavy or the sidewalk was icy, she would drive him to school, instead. Another family explained that wheelchair ramps were often too icy or snowy during winter to be useful. Other caregivers shared that they did not use the adapted school bus because it arrived too early to be useful for them in their busy mornings. So, sometimes, services meant to be accessible were not truly accessible for these families.

Evenings were described as being more relaxed than mornings. In the evenings, caregivers made dinner, had their children complete chores as they were able to, played games with their children, talked about school, and listened to audiobooks as a family. Caregivers took their children to specialized and frequent physical therapy and occupational therapy appointments. A few parents discussed taking time to cuddle with their younger children. School-aged children were expected to use some of the evening time for school work; a few parents noted their children’s academic and social successes:

> I’m proud of my kids. [Child with OI] does super well at school and has a nice personality.
>
> (Mother, Family 5, Type III)

Some children engaged in adapted sports, various artistic classes, and other low-impact activities. Older children with fewer accessibility challenges would go outside to play with friends on their own.

#### Facilitating mobilization

Mobilizing or transferring their children during these activities was conducted carefully. Some school-aged children would crawl or slide around on their buttocks, reserving walkers and wheelchairs for more fatiguing times. One child with a milder form of OI needed her family’s regular encouragement to move on her own, particularly up stairs where she had more difficulties. Transfers presented with some of the greatest difficulties for families. Many of the children needed at least some assistance in transferring to and from the bed, car, wheelchair, or toilet. One legal guardian shared that “just moving him from point a to point b is most challenging” (Guardian [Mother], Family 8, Type VI). While these caregivers typically lifted their school-aged child into the truck, it was becoming difficult to lift him as he was getting older and heavier. For families that had children with more severe types of OI, all of these activities facilitating mobilization needed to be accomplished without causing further pain or fractures. In contrast, families with Type I OI generally reported not noticing any significant differences in the day-to-day requirements of caring for their child living with OI, compared to caring for a child without OI.

#### Challenging periods

OI caregivers were constantly aware that the regular daily routines they had established could be disrupted by fractures, surgeries, and pain. These challenging periods increased the care management needed.

#### Fractures

The initial stress of bone breaks came from their unexpectedness. The first fractures in their child’s life caused some caregivers to panic, although caregivers said they learned to adapt. One mother shared that a challenging day for her was when she was called from her office due to a fracture at school:

> The challenging time is when I’m called from the office that there’s a fracture. Sometimes she fractures in school, sometimes she fractures when she comes back. And I will have to quickly rush home to go see the severity of the fracture, whether it’s just a crack, or it’s a real fracture where we have to go to the hospital. So basically, that’s when it becomes challenging.
>
> (Mother, Family 6, Type III)

She explained that she did not like going to the hospital because she said the hospitals in her country did not understand OI; she felt stigmatized in hospitals.

With more severe forms of OI, a fracture could happen practically spontaneously. One caregiver explained how easily femur fractures occurred:

> He was jumping on my bed. And, you know, like a normal four-year-old would do. We were doing laundry, he was jumping on the bed, and with that, we heard a *snap*. And he is *screaming*, leg bevelled out, and we’re like “well, that’s strange.” And then we went to the hospital, and they’re like, “okay, it’s a femur fracture.” And that happened about two other times after that. Then they’re like, “okay, we’re going to test him [for OI]”.
>
> (Guardian [Older Sister], Family 8, Type VI)

Another caregiver shared that whenever her child fractured, she immediately assessed the fracture and administered appropriate medications. Then, her child relaxed for the rest of day. Other parents shared that in the case of severe fractures, they would go to the hospital and wait for treatment and recovery.

#### Challenges of surgery

Surgeries presented unique difficulties for families. Hospital boredom was often a challenge for parents, especially if they had to fly to Montréal for surgery and wait several days or weeks before returning home. A few caregivers discussed how challenging planning their lives around surgeries could be – especially when, after arranging their work and family schedules around a specific date, the surgery date was sometimes changed by the hospital.

Care coordination for treatment created additional challenges for the three international families interviewed. Caregivers shared that accessing OI treatment in their own countries could be difficult, so a few of them received pro-bono treatment supplies through the international OI community. For a mother, whose daughter needed a few surgeries over several months, finding a Montreal sponsor to live with (as may sometimes be required) was a significant care coordination challenge for her. She said she had to somehow find a sponsor to live with for several months, in a city far from her own, while also arranging care with her husband and their other children who would stay behind in her home country. She explained:

> So I leave [the other children] with my husband when I came to the [Montreal OI treatment centre] in August … I think from the way it is, [the treatments] will end by December … You understand, I have my kids calling me, ‘Mommy, when are you coming back?’ … it’s a very long time.
>
> (Mother, Family 12, Type III)

#### Caregiving after a fracture or surgery

Caregiving became more difficult after a fracture or a surgery, with the child’s limb or limbs immobilized and unable to bear any weight. Immobility required more assistance with ADLs and made transfers more challenging. Leg fractures were particularly problematic because the child was then non-weight-bearing on that limb, making transfers to and from places - such as the bathtub, toilet, or car - heavy and complex for the caregiver. When children fractured, time for self-care and outside excursions could become more difficult:

> Um, especially when they’re fractured, it’s not as easy to get out. You don’t want to be transferring them when they’re in pain, getting them into their car-seat. And when you’re the only person at home ‘cause your husband’s working, if you need to go to the grocery store, then you have to take them with you. So sometimes you’re not necessarily getting yourself ready in the morning because you’re just trying to get them into a position where they can get out of the house at some point or for an appointment.
>
> (Mother, Family 13, Type III)

After surgery, the family’s routine would change. One mother explained that after a fracture, her child would sleep in the same bedroom as her, receive more pain medication, and stay home for a few days post-surgery. Fractures and surgeries created stress for caregivers during ADLs, as they were worried about any jostling during care work that could harm the child. As a result, caregivers tended to minimize activity and stay at home during these periods.

#### Pain

For some families of children with OI, life could revolve around pain management. One mother explained that when her school-aged son was in pain, he would call for “Mommy!” a lot more frequently (Mother, Family 5, Type III). Another mother shared that her child was often in physical pain, particularly experiencing regular headaches and back pain. As a caregiver, this mother had to evaluate the pain level two to three times per day and administer pain medications as needed. Two parents also shared that their children experienced regular back pain in spite of their fractures being healed. Periods of increased pain, then, created frequent disruptions to OI caregivers’ daily routines.

#### Long-term strategies for supporting day-to-day care

A comment by one mother embodied the management of day-to-day care:

> Dealing with OI is most likely all the time, how are we going to do things? You know? How are we going to surpass this limitation?
>
> (Mother, Family 5, Type III)

Caregivers became experts in caring for their children’s unique daily health and care needs. Caregivers developed strategies which would make future day-to-day caregiving work easier. They learned to manage the environment, access medical and school resources, and coordinate care and respite.

#### Managing the environment

Generally, caregivers tried to balance their desire to protect their child from harm, while also minimizing the limits placed on their child day-to-day. Managing care for caregivers often meant “managing the environment more than the kid”:

> We make sure that there is no chair or anything that could actually fall on her. the remote for the TV are never actually accessible if she’s standing from the couch, because … it’s heavy. It’s an old remote. We do play stuff differently, without saying “no you can’t” or “don’t do that” or “don’t touch that”. It’s really just managing the environment more than the kid … [The] environment is our responsibility.
>
> (Mother, Family 1, Type IV)

Managing the environment included adapting their housing to be safer (for instance, by installing extra-cushioned carpeting), coordinating with schools to manage the care of their child safely, and searching for low-impact activities that their children could participate in. With these environmental management strategies, future day-to-day care work was made easier, because their homes and schools were more prepared for a child with brittle bones to thrive in, and their children had low-impact activities and skills that they could develop.

#### Accessing medical and school supports

Caregivers described spending a lot of their caregiving time researching information and resources about OI, particularly for accessing medical and school supports. Once specialized medical and school supports were established, caregivers felt more at ease in their day-to-day care work.

##### Accessing medical support

For several caregivers, just because they had a diagnosis of OI did not mean they had adequate medical support and information for ongoing care. Caregivers shared that it was only once they found the Montreal OI treatment centre that they felt supported in their child’s medical care. Healthcare providers outside of the treatment centre – such as emergency staff and general pediatricians at other hospitals – did not always know how to treat OI. Some parents shared that when other health professionals tried to treat their children, the clinicians fractured their child’s other bones in the process. One Quebecoise mother believed not enough information was initially shared with her about her rights and available resources. She explained that when her daughter was a pre-schooler, the two of them spent an entire year avoiding all outings because they did not know how to mobilize her growing daughter beyond using a baby stroller. They were socially isolated and home-bound until she was told that her daughter had the right to access a properly fitted wheelchair.

Access to the greater OI community was an important resource for finding long-term medical supports. Several caregivers spent a lot of time seeking resources through their OI network. For instance, a few caregivers managed to receive regular OI treatment materials in their own country via the international OI community when they could not access those materials at home. Once caregivers managed to access long-term OI medical supports, either with an established OI social network or – ideally – with a specialized OI treatment centre nearby, caregivers felt more supported in their day-to-day caregiving activities, because they had access to people who were knowledgeable about OI; they were no longer caregiving alone.

##### Accessing school supports

For some families, finding daycare centres and schools that were comfortable with a child with OI was a challenging task. Once they found an accommodating school, several families described making plans to go into the school every year to share with the new educators how to care for a child with OI. These new school caregivers had to be taught:

> How do we immobilize a fracture, how do we give the medication, and how do we put him on the toilet seat … how do we take care of him?
>
> (Mother, Family 5, Type III)

Most children were attending public schools. However, they still required access to a school aide person who could help them in the washroom, carry books, push the wheelchair, or write for them, as needed.

A few caregivers were using more specialized school services. One child moved to Montreal with her mother to attend a school for children with special needs, while the child’s father and siblings remained in their hometown several hours away. Another family was choosing to homeschool their child instead of sending him to a public school, because they wanted to be sure he was mobilizing enough during the day. They feared that a public school would be too scared of him breaking a bone that they would not allow him to mobilize enough.

Other routine medical and school resource challenges included the availability of services, language barriers, and Internet access. Even among financially secure families, some families could not find appropriate daycare centres for their child with more severe OI. One mother explained that there was a long waiting list for daycare for children with special needs. Language barriers could also affect resource accessibility. Some families noted that the lack of OI resources available in languages outside of English (such as French) were limited. Access to the greater OI community through the Internet could also determine whether families managed to find a specialized OI treatment centre and other OI resources.

#### Coordinating care and accessing respite

Caregivers described many care coordination tasks related to their child’s OI. These included the following tasks: arranging and attending numerous medical appointments and treatments, remembering to bring the wheelchair or walker whenever they went out, managing respite care, being flexible with schedules in case of surgery or fractures, keeping track of medical records, seeking out community and health resources, and filling out medical paperwork and reimbursement forms. All of these activities required significant time and coordination skills on the part of caregivers, which frustrated some of the caregivers. However, once these coordination activities were accomplished, they made caregivers’ future day-to-day care work easier. For instance, by carefully organizing and keeping track of their child’s medical records, and by being flexible with their schedules, caregivers were able to quickly adapt if medical emergencies arose.

Most caregivers expressed chronic challenges with accessing respite care, explaining that they did not trust others to care for their child with OI. One mother said that learning to trust others is “sort of an issue” (Mother, Family 1, Type IV). Several caregivers shared that they were not comfortable hiring a babysitter for their child with OI, or even leaving their child with friends or family. Still, some families had grandparents and friends who caregivers were teaching how to care for their child in order to provide these caregivers with parenting breaks. Two mothers suggested that more trusted respite care opportunities, such as kids’ events or summer camps organized by OI treatment centres, would be very helpful. If caregivers had easy access to people they trusted to care for their child, day-to-day care was made easier because caregivers had a reliable back-up caregiver. However, most caregivers did not have anyone they fully trusted to provide this care.

## Discussion

This study offers a comprehensive overview of the day-to-day experiences of caring for a child with brittle bones, the challenging periods that can disrupt day-to-day routines, and the long-term strategies they developed to make day-to-day OI caregiving easier. Understanding the unique daily experiences of caregivers of children with rare and chronic conditions like OI is important knowledge for helping clinicians and policy makers to build better supports for these caregivers. Such supports could improve caregivers’ daily experiences of caregiving, and therefore improve their child’s care (Luijkx, Putten, & Vlaskamp, 2017; McCann et al., 2012).

The limited existing OI literature suggests that OI parents often have more tasks to accomplish in one day than most parents do. Even on typical days without fractures or surgery, a key difference is the child’s independence in completing ADLs. For more severe types of OI, in particular, the multiple physical transfers needed for toileting, clothing, and transport increased the energy and amount of time it took to accomplish these ADLs (Deatrick et al., 1988; Dogba et al., 2014). Facilitating mobilization and transportation for those living with rare and physical conditions is a common but challenging caregiving task (Andrews, 2013). However, in addition to managing the task of mobilization, OI caregivers of children with more severe types of OI also experience the unique and constant fear of fracturing their child while completing transfers and other day-to-day tasks (Brodin, 1993). Our study corroborates these reports of the experiences of OI day-to-day caregiving. In our study, caregivers shared that mornings were the most intense period of the day due to the number of ADLs needing attendance by a strict deadline (i.e., when school and caregivers’ paid work began). Furthermore, and in accordance with the research literature, facilitating their child’s mobilization and transportation was one of the most significant components of caregivers’ day-to-day routines.

For OI caregivers in our study, periods which presented particular challenges to OI caregivers’ day-to-day routines were periods with fractures, surgeries, and pain. These time periods were also described as some of the most challenging aspects of OI care in other studies of OI families (De Carmoy, 2004; Deatrick et al., 1988; Dogba et al., 2013; Santos et al., 2017). However, while the caregivers in our study acknowledged facing challenges and at times feeling overwhelmed, most appeared to be successfully managing as parents and caregivers. Caregivers explained that once they found the study site (a specialized OI treatment centre), they felt more supported as caregivers. These results correspond with the results of another study which was also conducted at our study site (Dogba et al., 2013). However, these results contrast with other studies on OI caregiving which described caregivers feeling burned out, hopeless, and overwhelmed by the unique challenges of caring for children with brittle bones (Arabaci et al., 2015; Bozkurt et al., 2014; De Carmoy, 2004). These latter studies do not appear to have been conducted in settings with access to specialized OI services. This contrast may speak to the psychosocial benefits that access to specialized OI medical resources can have on OI families.

The literature suggests that trying to manage the environment, access medical and school supports, coordinate care, and find trusted respite services are all common care management strategies of chronic caregivers (Pelentsov, Laws, & Esterman, 2015; Samia, O’Sullivan, Fallon, Aboueissa, & Hepburn, 2018). Other researchers have noted that OI caregivers spend significant amounts of time trying to gain reliable access to OI medical and school supports, social supports, and respite care (Arabaci et al., 2015; Bernehall & Brodin, 2002). These activities for facilitating day-to-day OI care over the long-term were described by caregivers in our study, as well.

## Strengths and Limitations

The methodological strengths of this study included providing rich descriptions of the study setting, participant demographics, and methods for transferability purposes. The results help address the paucity of literature available on OI caregiving, as well as on the supportive care needs of parents of children with a rare condition (Pelentsov et al., 2015). While corroborating much of the current OI caregiving literature, our study also adds to our knowledge of OI caregiving by providing more detailed data on what day-to-day OI caregiving looks like. This study appears to be the first to focus on the day-to-day tasks and routines of OI caregiving. Understanding the daily life of family caregivers is important because the differences required in care for caregivers of children with unique health needs can cause significant caregiver distress (Pelentsov et al., 2015). Furthermore, understanding their unique needs can support the development of evidence-based health and social supports for these families (Collins et al., 2016; Luijkx et al., 2017).

This study has several limitations. With the time and resources available for the study, only 18 caregivers from 14 families were interviewed. The sample could have benefited from more socioeconomic caregiver diversity. All caregivers had some post-secondary education, and a majority of the families (n=7) reported incomes of more than $80,000 per year. Only one caregiver was under the age of thirty, and all but two parents said they had another adult helping them at home, such as a spouse, adult child, or grandparent. The needs of younger and single-parent OI caregivers should be researched further. Finally, all of these families had access to the study site, which specializes in OI treatment. Researchers should explore the day-to-day experiences and routines of families who have more limited educational and financial resources, and who do not have access to a specialized centre for OI. Since the majority of the caregivers in this study had school-aged children (n=11 of the 17 children), more research should be conducted on the caregiving experiences of those caring for younger children, as well as those caring for teenagers. While these data revealed the day-to-day caregiving routines of regular days and challenging days, we did not fully explore what an excellent or optimal day looks like for caregivers of children with OI. Further qualitative and quantitative research should be conducted to build on this study’s work to better understand the diverse range of OI caregivers’ day-to-day caregiving experiences.

## Conclusion

In conclusion, the rich data collected from these interviews help elucidate the day-to-day activities and challenges faced by OI caregivers. The results complement previous research in the caregiving and OI research fields, while also adding a deeper understanding of the day-to-day experiences of OI caregivers. This knowledge may help clinicians and healthcare systems offer improved services to these families. It also provides further direction for future research initiatives to help promote the health and well-being of OI caregivers.

## Relevance to Clinical Practice

This study has implications for nurses and other pediatric clinicians. Finding information about OI, accessing treatment (especially for families who lived far away from the treatment center), finding knowledgeable local healthcare providers, and coordinating available community and school supports, were all challenges faced by the caregivers in our study. Clinicians should share the long-term strategies that these expert OI caregivers have developed, with newly-diagnosed families with OI. Doing so may help make the initial diagnosis less traumatic, giving families the practical information they need to care for their child at home in the community (Dogba et al., 2013). They should also work to develop clinical home-care tools to provide long-term supports for OI families. For instance, the study site has developed a splint kit that helps caregivers splint their children’s fractures at home; such a kit is particularly valuable when families do not have knowledgeable healthcare professionals nearby to assist them (***Will include citation once double-blind peer review is finished***). Similar resources could further support caregivers in helping their children to thrive with OI.

OI medical services should be more accessible. For instance, OI specialists should be easier to find. Nurses and clinical researchers should ensure that they are sharing accessible online resources for families coping with rare conditions like OI. For families coping with rare and relatively unknown conditions, the Internet may be one of their only sources of information (DeHoff, Staten, Rodgers, & Denne, 2016; Knapp, Madden, Wang, Sloyer, & Shenkman, 2011). OI caregivers shared that their day-to-day caregiving experiences were improved once they had access to the study site and specialized OI care. However, a few caregivers also shared that the time taken to travel to appointments, to complete surgeries, and to fill out reimbursement forms was too time-consuming and complicated. Institutions and policymakers should work to ensure that all medical services are accessible, efficient, and easy to navigate, with as little family separation as possible. Policymakers should focus on building supports for trusted respite care and other high-quality services. Policymakers should work to build more comprehensive and qualified respite support services for caregivers of care recipients with complex needs. A few caregivers in this study suggested that child care services provided by specialized treatment centres, such as special drop off days or summer camps, would be appreciated. Another option might be to develop OI caregiving teaching materials to help caregivers teach their friends and extended family on how to care for their child day-to-day. A few of the caregivers mentioned that even when they had community supports like adapted buses and wheelchair ramps, those services often did not meet their unique caregiving needs, so they did not use them. Policymakers should work to ensure that such services are adapted and truly accessible to families with complex care needs, so that these families can learn to trust and rely on the services developed for them.

## What does this paper contribute to the wider global clinical community?

- These results showcase what being a caregiver for a child with a rare and chronic physical condition - Osteogenesis Imperfecta - involves on a day-to-day basis.
- Caregivers of children with rare and chronic physical conditions like OI have unique needs, such as physical mobilization support needs, as well as needing better access to medical information, accessible schools, and appropriate care services.
- Nurses, clinicians, and policymakers who work with families coping with OI and similar conditions should develop interventions and policies that support the unique needs of these families; these interventions and policies should facilitate the long-term strategies that caregivers have already developed to support their day-to-day caregiving activities.

## Data Availability

Data was collected during interviews, not from any repositories.

## Acknowledgements

Several Shriners clinicians and caregiving stakeholders also provided invaluable feedback on the interview guides (Johanne Brunelle, RN; Sylvie-Anne Plourde, RN; Catherine Dubé, BSW; Trudy Wong, MSW; Mary Curtin, BA; Maria Caruso, BA, DipEd, CertSpEd; Marie Donato, BA, DipEd, CertEd; and Angela Gugliotti, BA). Finally, this study could not have been conducted without the caregiver participants who took time to share their knowledge and expertise.

